# Prediction of intensive care unit mortality based on missing events

**DOI:** 10.1101/2021.02.28.21252249

**Authors:** Tatsuma Shoji, Hiroshi Yonekura, Sato Yoshiharu, Yohei Kawasaki

## Abstract

**Background:** The increasing availability of electronic health records has made it possible to construct and implement models for predicting intensive care unit (ICU) mortality using machine learning. However, the algorithms used are not clearly described, and the performance of the model remains low owing to several missing values, which is unavoidable in big databases.

**Methods:** We developed an algorithm for subgrouping patients based on missing event patterns using the Philips eICU Research Institute (eRI) database as an example. The eRI database contains data associated with 200,859 ICU admissions from many hospitals (>400) and is freely available. We then constructed a model for each subgroup using random forest classifiers and integrated the models. Finally, we compared the performance of the integrated model with the Acute Physiology and Chronic Health Evaluation (APACHE) scoring system, one of the best known predictors of patient mortality, and the imputation approach-based model.

**Results:** Subgrouping and patient mortality prediction were separately performed on two groups: the sepsis group (the ICU admission diagnosis of which is sepsis) and the non-sepsis group (a complementary subset of the sepsis group). The subgrouping algorithm identified a unique, clinically interpretable missing event patterns and divided the sepsis and non-sepsis groups into five and seven subgroups, respectively. The integrated model, which comprises five models for the sepsis group or seven models for the non-sepsis group, greatly outperformed the APACHE IV or IVa, with an area under the receiver operating characteristic (AUROC) of 0.91 (95% confidence interval 0.89–0.92) compared with 0.79 (0.76–0.81) for the APACHE system in the sepsis group and an AUROC of 0.90 (0.89–0.91) compared with 0.86 (0.85–0.87) in the non-sepsis group. Moreover, our model outperformed the imputation approach-based model, which had an AUROC of 0.85 (0.83–0.87) and 0.87 (0.86–0.88) in the sepsis and non-sepsis groups, respectively.

**Conclusions:** We developed a method to predict patient mortality based on missing event patterns. Our method more accurately predicts patient mortality than others. Our results indicate that subgrouping, based on missing event patterns, instead of imputation is essential and effective for machine learning against patient heterogeneity.

**Trial registration:** Not applicable.

## Background

Accurate prognostication is central to medicine [1] and at the heart of clinical decision-making. Sepsis is a systemic response to infection, with the highest mortality rate in the field of intensive care. For nearly a decade, sepsis-related mortality has remained at 20%–30%, with unsatisfactory improvements [2]. Prognosis remains a challenge for physicians because of the high heterogeneity of clinical phenotypes [3]. Because accurate diagnoses can improve the physician’s decision-making abilities, it is one of the essence for medical practice of sepsis to improve prognosis accuracy [4].

Most large prognostic studies have developed clinical scoring systems for objective risk stratification in the early phase of hospital admission using physiological measurements, medical history, and demographics to predict the likelihood of survival [5]. Among these scoring systems, Acute Physiology and Chronic Health Evaluation (APACHE) scoring [6] is one of the best known, which has been validated for application at approximately 24 h after intensive care unit (ICU) admission. The APACHE scoring system generates a point score based on worst values of 12 variables during the initial 24 h after ICU admission. The APACHE II score was published in 1985 [5]; APACHE IV and IVa are the latest versions [7]. Built on the study of a more recent patient population and standard-of-care, APACHE IV or IVa are recommended as scoring systems over APACHE II and III. However, these indicators lack the precision required for use at the individual level. Therefore, efforts have been made to increase the performance of these indicators through the use of computational techniques, such as machine learning.

Machine learning classifiers may be advantageous for outcome prediction because they can handle large numbers of variables and learn non-linearities [8, 9]. Random forest is an example of modern machine learning algorithms [10]. Machine learning requires large volumes of data and generating a complex model. However, the increasing availability of electronic health records has made constructing and implementing the models possible. The availability of large training sets [11, 12] has made investigation of such approaches feasible. Studies on the application of machine learning to intensive care datasets have been performed [13, 14, 15, 16, 17, 18, 19, 20, 21]. Traditional scoring systems were outperformed by machine learning approaches in mortality prediction in medical ICUs. However, the algorithms are not clearly described and the produced models or databases are available commercially.

The Philips eICU Research Institute (eRI) is a non-profit institute established by Phillips that is governed by customers [22]. This freely accessible critical care dataset spans more than a decade and contains detailed information about individual patient care, including time-stamped, nurse-verified physiological measurements. As a distinctive feature, datasets are donated by >400, including teaching and non-teaching, hospitals. However, the eRI, which comprises 31 files according to clinical categories, includes much missing data. This makes the application of machine learning with eRI challenging.

In this study, we revealed unique characteristics of the distribution of missing values in the eRI database. By taking advantage of this characteristic, we successfully developed a more accurate and interpretable model against the APACHE system. Accurate prediction of patient state is critical in the critical care field. To address this problem, we propose the “missing-event-based prediction,” a new method for predicting ICU mortality.

## Methods

### Dataset used in this study

We used the eRI (https://www.usa.philips.com/healthcare/solutions/enterprise-telehealth/eri) database, which is an open database and provides anonymous data, including demographic information, vital signs measurements, laboratory test results, drug information, procedural information, fluid balance reports, hospital length of stay data, and data on in-hospital mortality, donated by >400 member institutions. The eRI contains data associated with 200859 ICU admissions with more than 100 variables. In this study, we carefully selected 58 clinically important variables to construct a model, which is easy to interpret clinically. Selected variables are summarized in Table S1 (see Additional file 1). Briefly, the variables were as follows: (I) Listed from laboratory measurements, including white blood cell count, hematocrit, bilirubin, creatinine, sodium, albumin, blood urea nitrogen, glucose, arterial pH, fraction of inspired oxygen, arterial oxygen pressure, and arterial blood carbon dioxide pressure. (II) Listed from routine charted data, including temperature, respiratory rate, heart rate, mean arterial blood pressure, urine output, and Glasgow Coma Scale (including score for eye, motor, and verbal responses). (III) Listed from information taken at the time of ICU admission, including age, gender, height, weight, time from hospitalization to ICU admission, type of hospital, bed count of the hospital, hospital ID, and diagnoses names at ICU admission. (IV) Comorbidities, including myocardial infarction within 6 months, diabetes, hepatic failure, dialysis, immunosuppressive disease, lymphoma, leukemia, metastatic cancer, cirrhosis, acquired immune deficiency syndrome, and history of intubation and mechanical ventilation. Intervention required at admission, including catheter intervention for myocardial infarction, coronary artery bypass grafting with or without internal thoracic artery grafts, and use of thrombolytics. (V) APACHE scoring system, including not only the APACHE score but also actual ICU and in-hospital mortality, predicted ICU and in-hospital mortality, length of ICU stay, length of hospital stay, and ventilation duration. Among these variables, diagnoses names at ICU admission were used to define the sepsis or non-sepsis group (see the Definition of sepsis and non-sepsis groups in Methods section for details), actual ICU and in-hospital mortality were used as response variables to construct the models, predicted ICU and in-hospital mortality using the APACHE IV or IVa were used as benchmarks against our model, and others (53 parameters in total) were used as explanatory variables for machine learning.

### Definition of sepsis and non-sepsis groups

Inclusion criteria for the sepsis group were as follows: (I) extraction by diagnoses names at ICU admission, namely “Sepsis, cutaneous/soft tissue,” “Sepsis, GI,” “Sepsis, gynecologic,” “Sepsis, other,” “Sepsis, pulmonary,” “Sepsis, renal/UTI (including bladder),” and “Sepsis, unknown;” (II) selection by documentation of prognosis; and (III) exclusion by cases with any missing data in Acute Physiology Score (APS)-related variables and prognosis information. Inclusion criteria for the non-sepsis group were almost the same as the one used for the sepsis group. Briefly, the complementary subset of (I) was first selected. Then, (II) and (III) were applied to the resulting subset. Thus, 4226 and 23170 cases were defined as sepsis and non-sepsis groups, respectively. To reproduce our results and access patient lists, follow the jupyter notebook at https://github.com/tatsumashoji/ICU/1_the_sepsis_group_and_non_sepsis_group.ipynb.

### Subgrouping based on missing data

Subgroups were defined according to the diagram shown in Fig. S1 (see Additional file 2). Briefly, (I) patient lists, containing no missing data for any pattern of 52 variables (derived from 53 variables) were first generated. Then, (II) the patient list, which had a size not too small and not too large among 53 lists, was defined as subgroup #1. For the other subgroups, we repeated (I) and (II) with the other patients. To reproduce this subgrouping, follow the jupyter notebook opened at https://github.com/tatsumashoji/ICU/2_subgrouping_sepsis.ipynb for the sepsis group and https://github.com/tatsumashoji/ICU/3_subgrouping_non_sepsis.ipynb for the non-sepsis group.

### Generation and performance of our model

To construct the model for each group, we used the random forest classifier implemented with “scikit-learn (0.24.1)” [23]. Briefly, we first selected 80% data as a training dataset for each group so that the ratio of “ALIVE” and “EXPIRED” cases were the same between the two datasets. After hyperparameters for the random forest were determined using the grid search algorithm, the actual model was generated, and the mean and standard deviation of accuracy were checked through 5-fold cross-validation (see Table S2-S5 in Additional file 1). Finally, patient mortalities in the test dataset were predicted by the generated model and compared to those from APACHE IV or IVa by drawing receiver operating characteristic (ROC) curves and calculating the area under the ROC (AUROC). The confidence intervals for the AUROC were calculated as described [24]. For calibration plots, we used the module “sklearn.calibration.calibration_curve.” All results in Fig. 2 can be reproduced by running “https://github.com/tatsumashoji/ICU/4_sepsis_prediction.ipynb.”

### Imputation of missing values

For imputation of missing values, we used multivariate imputation algorithms implemented with “sklearn.impute.IterativeImputer,” which uses the entire set of available feature dimensions to estimate missing values. We used the 0.24.1 version of scikit-learn. To reproduce results shown in Fig. 3, follow the jupyter notebook opened at https://github.com/tatsumashoji/ICU/5_imputation.ipynb.

## Results

### Heterogeneity of the sepsis group

Machine learning fails to predict the outcomes if input data consist of more than two populations. Subgrouping of input data is essential before constructing the model using machine learning. Therefore, we first investigated the distribution of each parameter in the sepsis group. The histogram in Fig. 1a shows the distribution of some variables recorded in “apacheApsVar.csv”, which contains the variables used to calculate the Acute Physiology Score (APS) III for patients. More than two peaks were observed, indicating the necessity for subgrouping the sepsis group. Importantly, cases with missing data on any variable are excluded from the histograms in Fig. 1a. Thus, to examine how missing values were distributed in the sepsis group, correlation coefficients were calculated for all possible combinations of two variables after missing data were replaced with 0 and others with 1 (Fig. 1b). Surprisingly, perfect correlations were observed in some pairs in addition to the diagonal line. This suggests that missing events occur depending on other missing events. Clinically, this can be explained by different histories and backgrounds in the sepsis group. Thus, missing events contain information and may play an important role in subgrouping the sepsis group.

**Fig 1.**
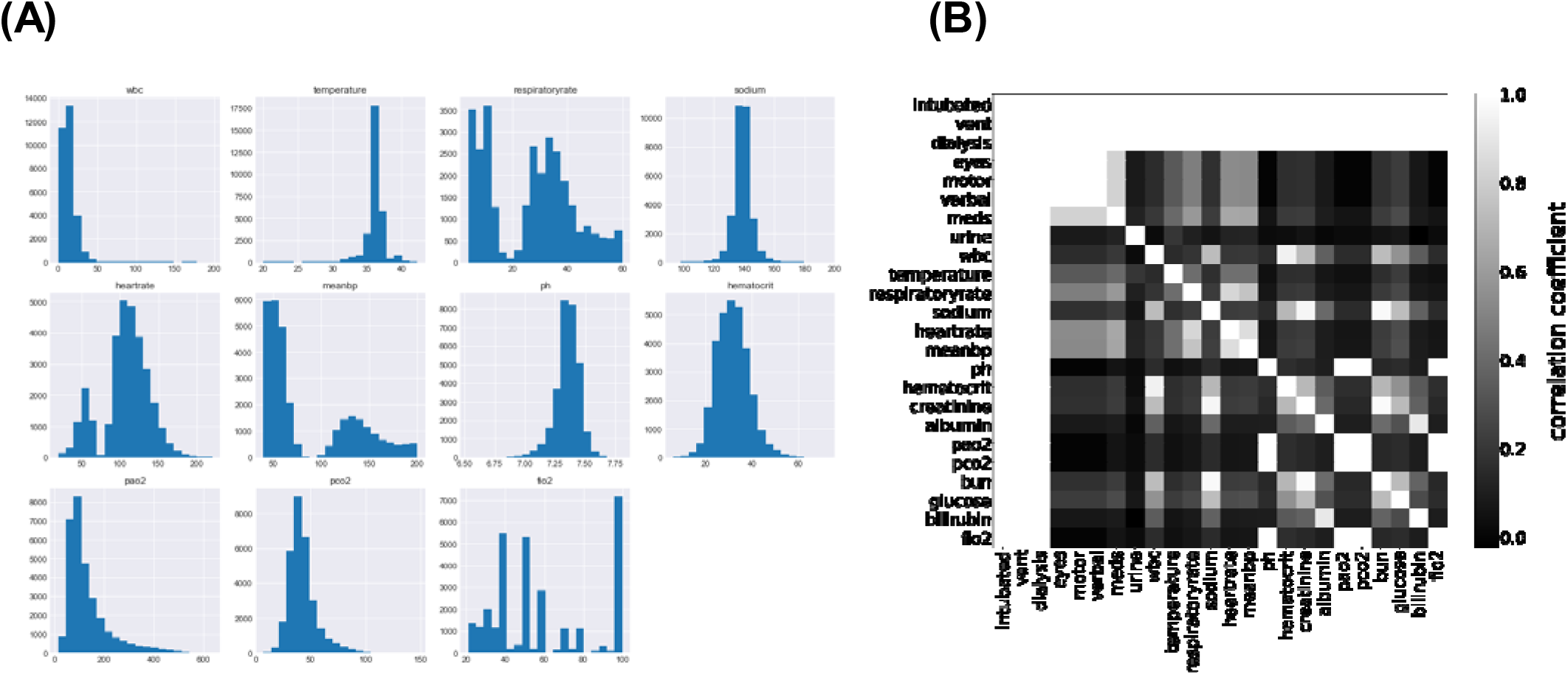
Distribution of Acute Physiology Score (APS)-related variables. **a** The distribution of APS-related variables in the sepsis group. **b** Joint matrix for checking the missing values. Correlation coefficients in some combinations were picked up and visualized using the heatmap where higher coefficients were lighter and lower were darker.

### Machine learning combining missing-event-based subgrouping approach outperforms APACHE

The unique distribution of missing values in the sepsis group led us to divide the group before machine learning. To account for the pattern of missing events when dividing the sepsis group, we defined subgroups such that each subgroup had the same missing pattern while the number of subgroups could as small as possible and the size of each subgroup could be as large as possible (Fig. 2a). For details, see the Methods section. After defining five subgroups, we constructed models for each subgroup based on the random forest algorithm and calculated patient mortality. Then, we assessed the performance of each model by calculating AUROC and compared them to those from the APACHE IV or IVa system (Fig. 2b). Our model outperformed the APACHE systems, especially when integrating subgroups. Moreover, our model was more successful that APACHE in distinguishing patient mortality (Fig. 2c). Furthermore, results from calibration plots supported the ability of our model to predict patient mortality (Fig. 2d). Our model tends to output patient mortality higher than actual, whereas the APACHE system does not, indicating that the APACHE system underestimates patients’ mortality.

**Fig 2.**
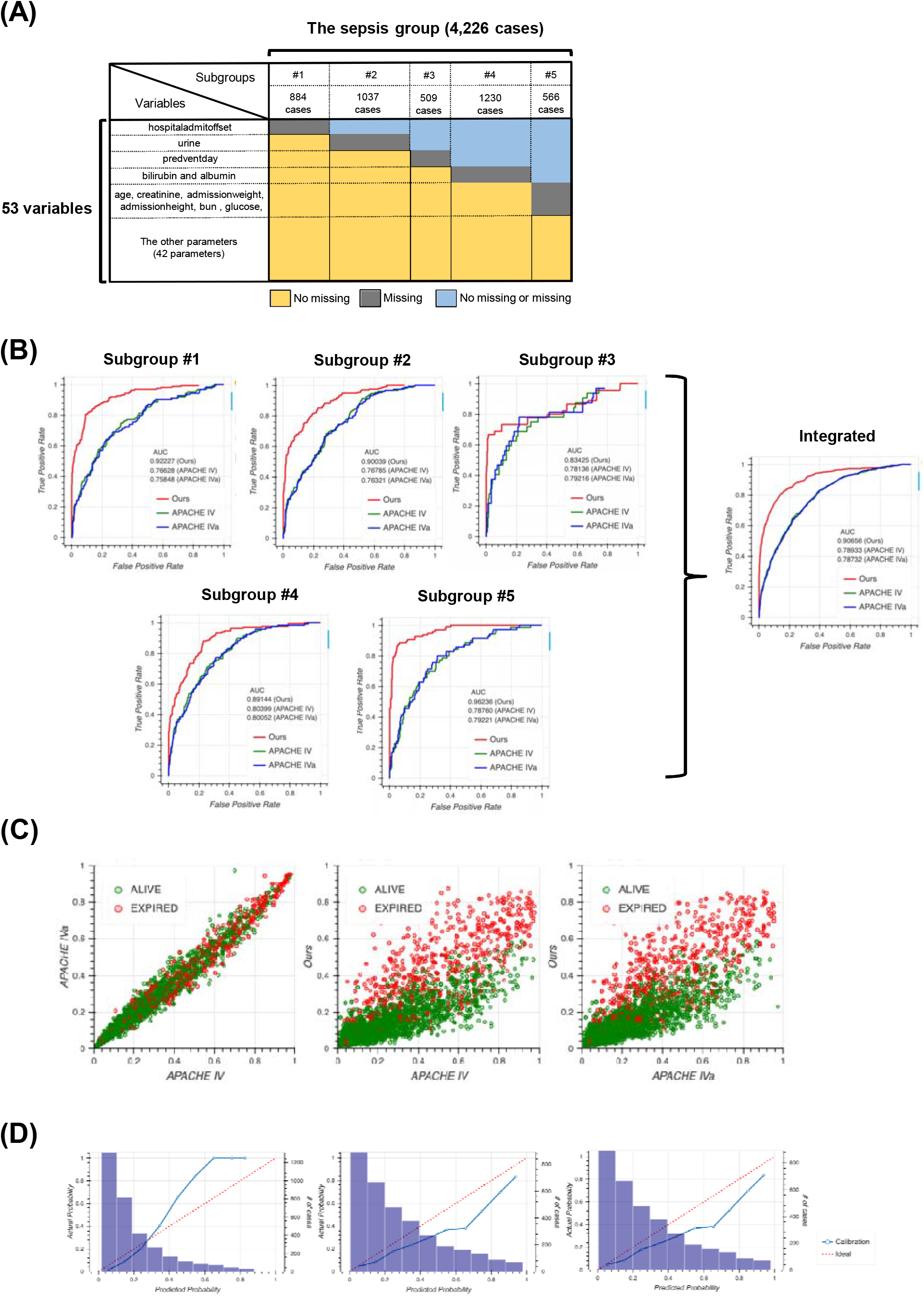
The performance of our model. **a** Schematic of subgrouping results. The sepsis group was divided into five subgroups, namely subgroup #1 (884 cases), #2 (1037 cases), #3 (509 cases), #4 (1230 cases), and #5 (566 cases). Colors indicate information about missing values. For example, cases in subgroup #1 have no missing values for all variables except “hospitaladmitoffset”. **b** Receiver operating characteristic (ROC) curves for each subgroup produced using our model (red), Acute Physiology and Chronic Health Evaluation (APACHE) IV (green), and APACHE IVa (blue). The integrated version is shown on extreme right. For the confidence interval of AUROC, see Table S6 in Additional file 1. **c** Scatter plots of predicted mortality; APACHE IV vs. APACHE IVa (left), APACHE IV vs. our model (middle), and APACHE IVa vs. our model (right). Colors indicate actual mortality (green for “ALIVE” and red for “EXPIRED” cases). For scatter plots of each subgroup, see Fig. S2 in Additional file 2. **d** Calibration plots for our model (left), APACHE IV (middle), and APACHE IVa (right). For the plot for each subgroup, see Fig. S3 in Additional file 2.

### Comparison with the imputation approach

A typical way of handling missing data is to impute them. Therefore, we constructed the model completely same as the way taken in Fig. 2 after imputing the dataset first, then compared to our model. Although the performance when imputed was slightly higher than the APACHE systems, it still remained lower than our method (Fig. 3a), indicating that missing events are important information and our method, which accounts for the pattern of missing events, is reasonable. The imputation approach outperforms the approach of the APACHE system but does perform as well as our model, as confirmed using the scatter plot (Fig. 3b). Our model distinguished patient mortality most precisely among all four models. Moreover, analyses of calibration plots supported the observation that our model is the most conservative, and thus, safer, because the model based on the imputation approach estimated lower patient mortality compared with our model (Fig. 3c).

**Fig 3.**
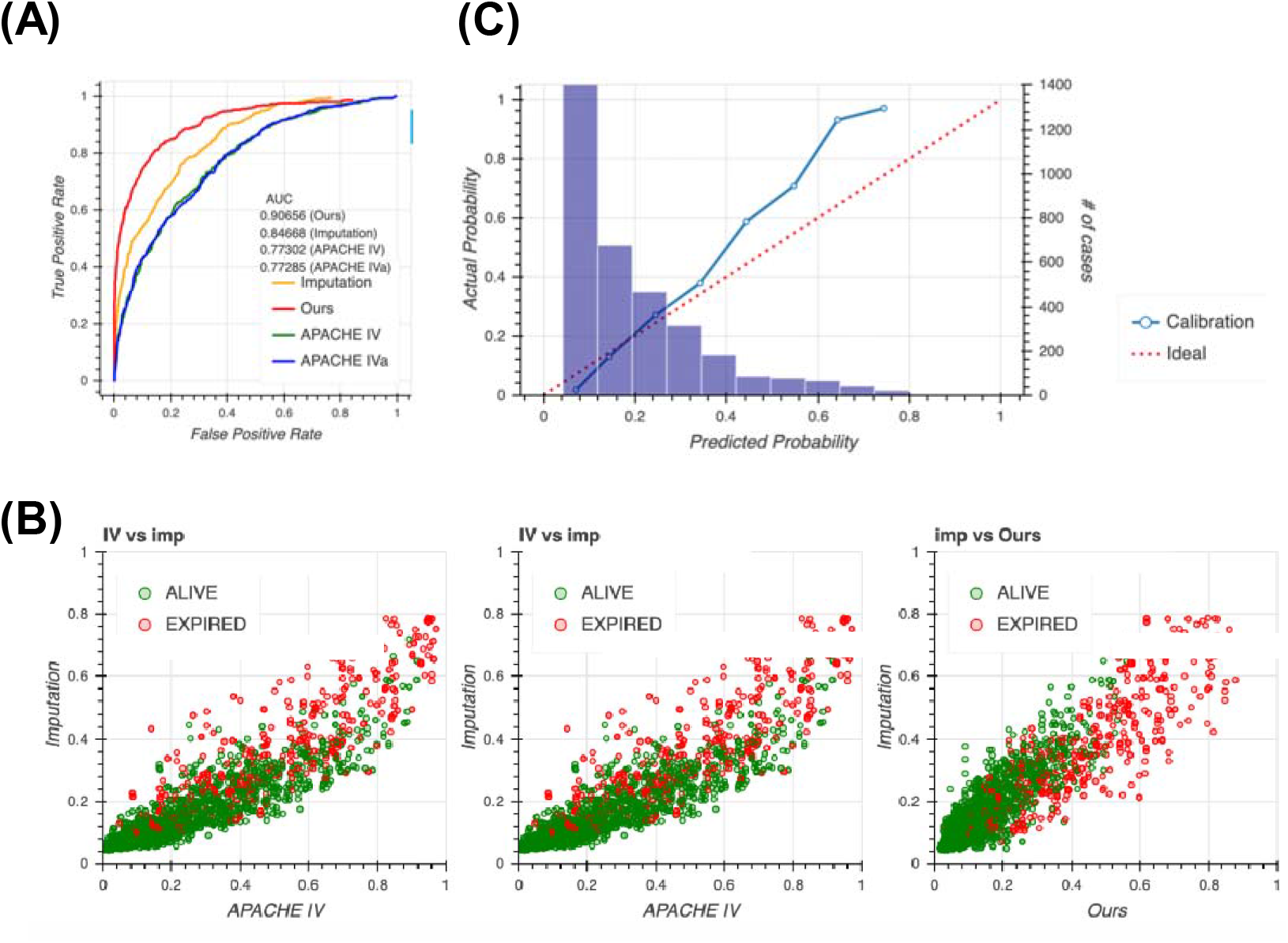
Performance of our model against the model based on the imputation approach. **a** Receiver operating characteristic (ROC) curves for the four models, namely our model (red), Acute Physiology and Chronic Health Evaluation (APACHE) IV (green), APACHE IVa (blue), and imputation (orange). For the confidence interval of AUROC, see Table S7 in Additional file 1. **b** Scatter plots of predicted mortality; imputation vs. APACHE IV (left), imputation vs. APACHE IVa (middle), and imputation vs. our model (right). Colors indicate actual mortality (green for “ALIVE” and red for “EXPIRED” cases). **c** Calibration plots for the model based on the imputation approach.

### Application to the non-sepsis group

To test the generalizability of our model, we applied our method to the non-sepsis group defined in the Methods section. We first generated seven subgroups using the same algorithm used for the sepsis group (Fig. 4a) and constructed models for each subgroup. Then, we compared the performance of the four models, namely our model, APACHE IV, APACHE IVa, and the model based on the imputation approach (Fig. 4b). Surprisingly, the distribution of missing values in the non-sepsis group was almost the same as that in the sepsis group, indicating that our subgrouping algorithm could be used for any group in addition to the sepsis group, and the performance of our model was the highest among all four models. Scatter plots and calibration plots also supported our model (Fig. S4, S5, see Additional file 2). These results strongly suggest that missing events themselves are essential when predicting patient mortality in the ICU.

**Fig 4.**
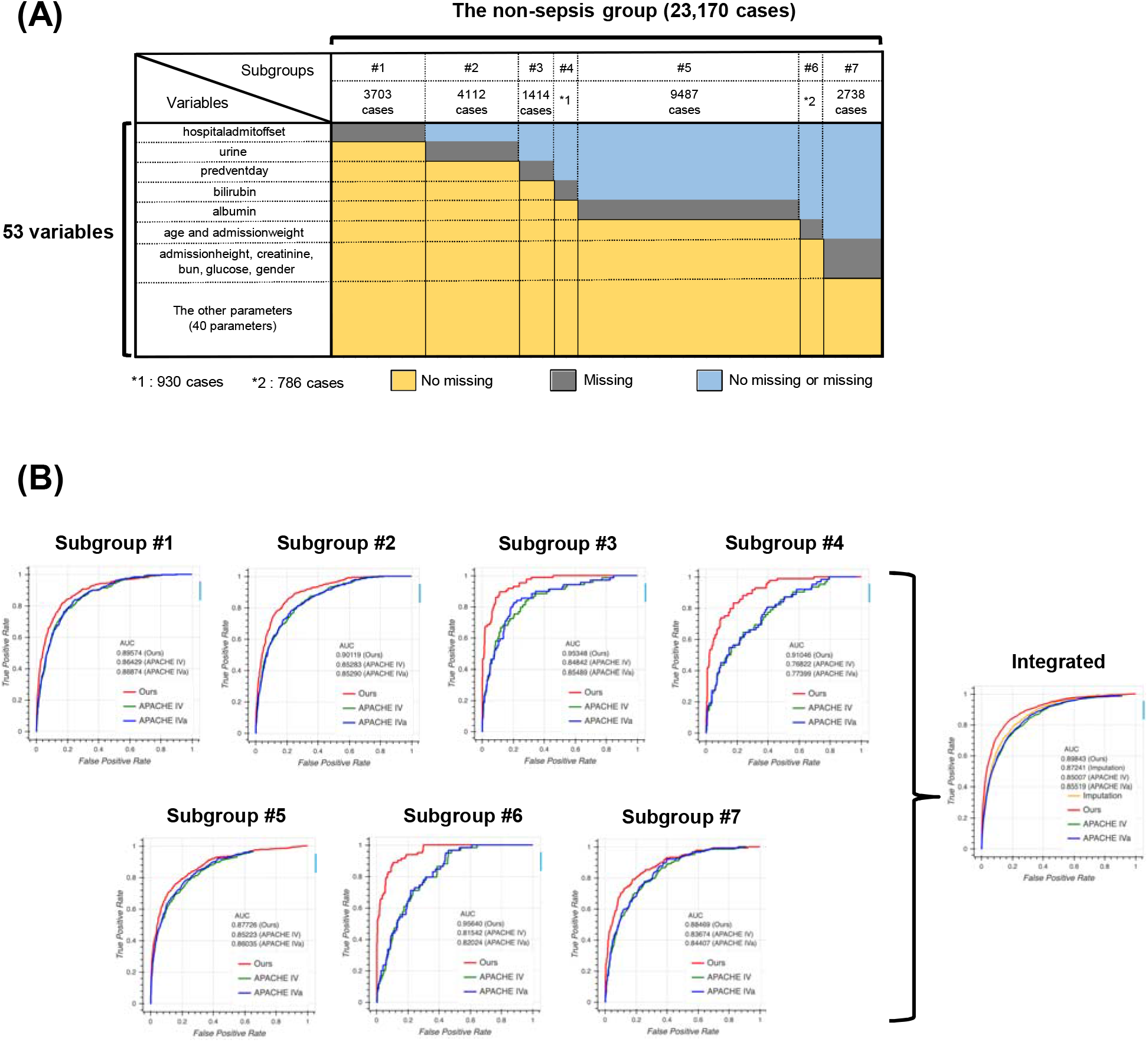
Performance in the non-sepsis group. **a** Schematic image for the result of subgrouping. The non-sepsis group was divided into seven subgroups, namely #1 (3703 cases), #2 (4112 cases), #3 (1414 cases), #4 (930 cases), #5 (9487 cases), #6 (786 cases), and #7 (2738 cases). Colors indicate the information about missing cases. For example, cases in the subgroup #1 have no missing values for all variables except “hospitaladmitoffset”. **b** Receiver operating characteristic (ROC) curve for each subgroup using our model (red), Acute Physiology and Chronic Health Evaluation (APACHE) IV (green), and APACHE IVa (blue). The integrated version is shown on extreme right with the result from the imputation approach. For the confidence interval of the area under the ROC (AUROC), see Table S8 in Additional file 1.

## Discussion

The main objective of this work was to present a more accurate and clinically interpretable model for predicting patient mortality in the ICU and show the effectiveness and potential of the missing-event-based prediction method. Our analysis of the eRI confirmed that the larger the database across hospitals, the more the heterogeneity (Fig. 1a). This is known as the domain shift problem, where traditional models, including the APACHE system, developed in one geographical region or healthcare system lose their ability to discriminate when applied outside of learning data. The effect of prediction accuracy by volume and diversity of missing events clearly increases in big data. We demonstrated the advantage of using a missing-event-based method through comprehensive analysis and presented a more accurate and interpretable model than others.

In the eRI database, missing events occurred depending on other missing events (Fig. 1b). Thus, missing events were not random. Such cases would not be rare in big data because the larger the dataset, the more complex the missing events. If missing event was not regarded “missing at random,” the simple imputation of missing values would not be appropriate [25]. For example, Meiring et al. developed an algorithm to predict mortality over time in the ICU using CCHIC (a UK database) [20]. They imputed missing values using predictive mean matching through parallel implementation of multiple imputation by chained equations [26]. In this report, the discriminative power of the APACHE II score to predict outcomes on subsequent days reduced considerably. Moreover, Meiring et al. indicated that the longer the ICU stay, the smaller the size of the dataset, and therefore, an increase in the proportion of missing values. Thus, the weight of missing values impaired the accuracy of prediction. Handling missing values is important to generate a powerful model workable on complex clinical courses. Thus, our missing-event-based method is reasonable. Because the subgrouping pattern in this study was humanly impossible to detect, although easy to understand, our subgrouping algorithm could be the new *de facto* standard of database prescreening when constructing an accurate and interpretable model through machine learning using big data, including missing values. Our subgrouping algorithm can recognize missing events as an “informative missingness” instead of a dirty record, which big data cannot avoid including.

For constructing the model based on the random forest algorithm, we selected 53 variables as explanatory variables so that they were closely related to the APACHE system. This selection is an important aspect of the interpretability of our model because the APACHE system consists of clinically used variables. This feature is quite important because the interpretable model addresses the problem of the “black box”, which has hindered the use of this model as a clinical tool [27, 28, 29]. Given that our model greatly outperformed the APACHE system, it can be considered a developed version of the APACHE system, which detects the presence of more complex interactions between covariates, leading to optimization both of clinical usability and discrimination ability.

As tested with the non-sepsis group, our method is applicable to other cases, indicating that it may not depend on diagnoses in the ICU. Under intensive care conditions, avoiding the occurrence of missing events is a challenge; therefore, our method can be useful in the clinic. Distinguish a generalizable method from a generalizable predictive model for clinical applications of machine learning is important. This study shows availability of subgrouping based on missing events as a generalizable method and production of a promising predictor model as a clinical decision support tool.

## Conclusions

We developed a method to predict patient mortality based on information on missing events. This method more accurately predicted patient mortality than others, while maintaining clinical interpretability. Our results indicate that the subgrouping process is important and effective for machine learning against patient heterogeneity. By combining our method with other methods, such as the reinforcement learning, a more realistic Artificial Intelligence clinician can be developed.

## Data Availability

The datasets generated and/or analyzed during the current study are available in the eICU repository.

https://eicu-crd.mit.edu/gettingstarted/access/

## List of abbreviations

APACHE: Acute Physiology and Chronic Health Evaluation
ICU: Intensive Care Unit
eRI: eICU Research Institute
APS: Acute Physiology Score
ROC: Receiver Operating Characteristic
AUROC: Area Under the ROC

## Declarations

### Ethics approval and consent to participate

Not applicable.

## Consent for publication

Not applicable.

## Availability of data and materials

The datasets generated and/or analyzed during the current study are available in the eICU repository, [https://eicu-crd.mit.edu/gettingstarted/access/].

## Author contributions

ST analyzed the eRI data, suggested, and implemented subgrouping algorithm and random forest model, prepared all of Figures and Tables and was a major contributor in writing the manuscript. YH selected the 53 explanatory variables in the point of clinical view, was the other major contributor in writing the manuscript as a clinician and supported the research grant. SY checked the programs that ST wrote, reviewed the manuscript and supported the research grant. KY contributed to the conception and overall design of the work, reviewed the results and manuscripts as a clinical statistician and supported the research grant. Finally, all four authors played an essential role in organizing this project.

## Acknowledgments

We thank Ryota Jin for discussions and comments to the manuscript.

## Supplementary Information

### Additional file 1

File name: Additional_file_1.xlsx

File format: Microsoft excel.

Title of data: Tables S1–S8.

Description of data: Supplementary tables.

### Additional file 2

File name: Additional_file_2.pptx File format: Microsoft power point. Title of data: Figures S1–S5.

Description of data: Supplementary figures.

## Supplementary Figure Legends

**Fig. S1 Flowchart for generating subgroups**.

The blue box at the top indicates the starting point of the flowchart. Subgroups for the sepsis or non-sepsis group are generated by following steps of the flowchart. The set *P* is the defined patient list (*P* = 4226 for the sepsis group and *P* = 23170 for the non-sepsis group). *V* is the set of explanatory variables (i.e., the size of the *V* is 53). *N*_*min*_ and *N*_*max*_ are set at 500 and 2000, respectively, for the sepsis group and 500 and 10000, respectively, for the non-sepsis group. 2^*V*^ denotes the power set of set *V. V*_*dj*_^*c*^ denotes the complementary subset for *V*_*dj*_. n(*A*) denotes the size of set *A*.

**Fig. S2 Scatter plot for the predicted mortality of each subgroup**.

Scatter plots for predicted mortality; Acute Physiology and Chronic Health Evaluation (APACHE) IV vs. APACHE IVa (left column), APACHE IV vs. our model (middle column), and APACHE IVa vs. our model (right column). Colors indicate actual mortality (green for “ALIVE” and red for “EXPIRED” cases).

**Fig. S3 Calibration plots for each subgroup**.

Calibration plots for our model (left column), Acute Physiology and Chronic Health Evaluation (APACHE) IV (middle column), and APACHE IVa (right column).

**Fig. S4 Scatter plots of predicted mortality in the non-sepsis group**.

Scatter plots of predicted mortality for each subgroup and integrated version; Acute Physiology and Chronic Health Evaluation (APACHE) IV vs. APACHE IVa (left column), APACHE IV vs. our model (middle column), and APACHE IVa vs. our model (right column). Three panels at the bottom show the comparison with the model generated based on the imputation approach. Colors indicate actual mortality (green for “ALIVE” and red for “EXPIRED” cases).

**Fig. S5 Calibration plots in the non-sepsis group**.

Calibration plots in the non-sepsis group for each subgroup and the integrated version. Columns on the left, middle, and right show results from our model, Acute Physiology and Chronic Health Evaluation (APACHE) IV, and APACHE IVa, respectively. The bottom panel shows results using the model generated based on the imputation approach.

## Supplementary Table Legends

**Table S1 Working dataset used in this study**.

The column on the left corresponds to file names, which can be downloaded from the eRI. We used 4 out of 31 files. The column in the middle shows the names of variables selected in this study. The column on the right indicates the role of variables in this study, where “key” indicates the unique key used for merging each .csv file and “x” and “y” indicate explanatory and response variables for machine learning, respectively, Acute Physiology and Chronic Health Evaluation (APACHE) indicates predicting patient mortality using APACHE IV or IVa, which was used as a benchmark against our model, and “classification” indicates the variable that is used to define the sepsis or non-sepsis group.

**Table S2 Results from cross-validation for each subgroup from the sepsis group**.

**Table S3 Results from cross-validation for imputed data from the sepsis group**.

**Table S4 Results from cross-validation for each subgroup from the non-sepsis group**.

**Table S5 Results from cross-validation for imputed data from the non-sepsis group**.

**Table S6 Confidence interval of area under the receiver operating characteristic (AUROC) for each subgroup and the integrated version**.

**Table S7 Confidence interval of area under the receiver operating characteristic (AUROC) for all four models**.

**Table S8 Confidence interval of area under the receiver operating characteristic (AUROC) for each subgroup and the integrated version in the non-sepsis group**.

## References

1. Moons KG, Royston P, Vergouwe Y, Grobbee DE, Altman DG. Prognosis and prognostic research: what, why, and how? BMJ. 2009;338:b375.

2. Rudd KE, Johnson SC, Agesa KM, Shackelford KA, Tsoi D, Kievlan DR, et al. Global, regional, and national sepsis incidence and mortality, 1990–2017: analysis for the Global Burden of Disease Study. Lancet. 2020;395:200–11.

3. Seymour CW, Kennedy JN, Wang S, Chang CC, Elliott CF, Xu Z, et al. Derivation, validation, and potential treatment implications of novel clinical phenotypes for sepsis, J Am Med Assoc. 2019;321:2003.

4. Schinkel M, Paranjape K, Nannan Panday RS, Skyttberg N, Nanayakkara PWB. Clinical applications of artificial intelligence in sepsis: a narrative review. Comput Biol Med. 2019;115:103488.

5. Vincent JL, Moreno R. Clinical review: scoring systems in the critically ill. Crit Care. 2010;14:207.

6. Knaus WA, Draper EA, Wagner DP, Zimmerman JE. APACHE II: a severity of disease classification system. Crit Care Med. 1985;13:818–29.

7. Zimmerman JE1, Kramer AA. Outcome prediction in critical care: the Acute Physiology and Chronic Health Evaluation models. Curr Opin Crit Care. 2008;14:491–7.

8. Deo RC. Machine learning in medicine. Circulation. 2015;132:1920–30.

9. Kourou K, Exarchos TP, Exarchos KP, Karamouzis MV, Fotiadis DI. Machine learning applications in cancer prognosis and prediction. Comput Struct Biotechnol J. 2015;13:8–17.

10. Breiman L. Random forests. Mach Learn. 2001;45:5–32.

11. Johnson AE, Pollard TJ, Shen L, Li-wei HL, Feng M, Ghassemi M, et al. MIMIC-III, a freely accessible critical care database. Sci Data. 2016;3:160035.

12. Harris S, Shi S, Brealey D, MacCallum NS, Denaxas S, Perez-Suarez D, et al. Critical Care Health Infor-matics Collaborative (CCHIC): data, tools and methods for reproducible research: a multi-centre UK intensive care database. Int J Med Inform. 2018;112:82–9.

13. Dybowski R, Weller P, Chang R, Gant V. Prediction of outcome in critically ill patients using artificial neural network synthesised by genetic algorithm. Lancet. 1996;347:1146–50.

14. Jaimes F1, Farbiarz J, Alvarez D, Martínez C. Comparison between logistic regression and neural networks to predict death in patients with suspected sepsis in the emergency room. Crit Care. 2005;9:R150–6.

15. Calvert J, Mao Q, Hoffman JL, Jay M, Desautels T, Mohamadlou H, et al. Using electronic health record collected clinical variables to predict medical intensive care unit mortality. Ann Med Surg (Lond). 2016;11:52–7.

16. Taylor RA, Pare JR, Venkatesh AK, Mowafi H, Melnick ER, Fleischman W, et al. Prediction of in-hospital mortality in emergency department patients with sepsis: a local big data-driven, machine learning approach. Acad Emerg Med. 2016;23:269–78.

17. Calvert J, Mao Q, Hoffman JL, Jay M, Desautels T, Mohamadlou H, et al. Using electronic health record collected clinical variables to predict medical intensive care unit mortality. Ann Med Surg (Lond). 2016;11:52–7.

18. Ward L, Paul M, Andreassen S. Automatic learning of mortality in a CPN model of the systemic inflammatory response syndrome. Math Biosci. 2017;284:12–20.

19. Aushev A, Ripoll VR, Vellido A, Aletti F, Pinto BB, Herpain A, et al. Feature selection for the accurate prediction of septic and cardiogenic shock ICU mortality in the acute phase. PLoS One. 2018;13:e0199089.

20. Meiring C, Dixit A, Harris S, MacCallum NS, Brealey DA, Watkinson PJ, et al. Optimal intensive care outcome prediction over time using machine learning. PLoS One. 2018;13:e0206862.

21. García-Gallo JE, Fonseca-Ruiz NJ, Celi LA, Duitama-Muñoz JF. A machine learning-based model for 1-year mortality prediction in patients admitted to an Intensive Care Unit with a diagnosis of sepsis. Med Intensiva. 2020;44:160–70.

22. Pollard TJ, Johnson AE, Raffa JD, Celi LA, Mark RG, Badawi O. The eICU Collaborative Research Database, a freely available multi-center database for critical care research. Sci Data. 2018;5:1–13.

23. Pedregosa F, Varoquaux G, Gramfort A, Michel V, Thirion B, Grisel O, et al. Scikit-learn: machine learning in python. J Mach Learn Res. 2011;12:2825–30.

24. Cortes C, Mohri M. Confidence intervals for the area under the ROC curve. Adv Neural Inform Process Syst. 2015;17:305–12.

25. Mallinckrod CH, Lane PW, Schnell D, Peng Y, Mancuso JP. Recommendations for the primary analysis of continuous endpoints in longitudinal clinical trials. Drug Inf J. 2008;42:303–19.

26. Buuren SV, Groothuis-Oudshoorn K. mice: multivariate imputation by chained equations in R. J Stat Softw. 2011;45:1–67.

27. Calvert JS, Price DA, Chettipally UK, Barton CW, Feldman MD, Hoffman JL, et al. A computational approach to early sepsis detection. Comput Biol Med. 2016;74:69–73.

28. Calvert J, Desautels T, Chettipally U, Barton C, Hoffman J, Jay M, et al. High-performance detection and early prediction of septic shock for alcohol-use disorder patients. Ann Med Surg. 2016;8:50–5.

29. Desautels T, Calvert J, Hoffman J, Jay M, Kerem Y, Shieh L, et al. Prediction of sepsis in the intensive care unit with minimal electronic health record data: a machine learning approach. JMIR Medical Informatics. 2016;4:e5909.

