## Additional file 2 for "Prediction of intensive care unit mortality based on missing events"

### Slide 1
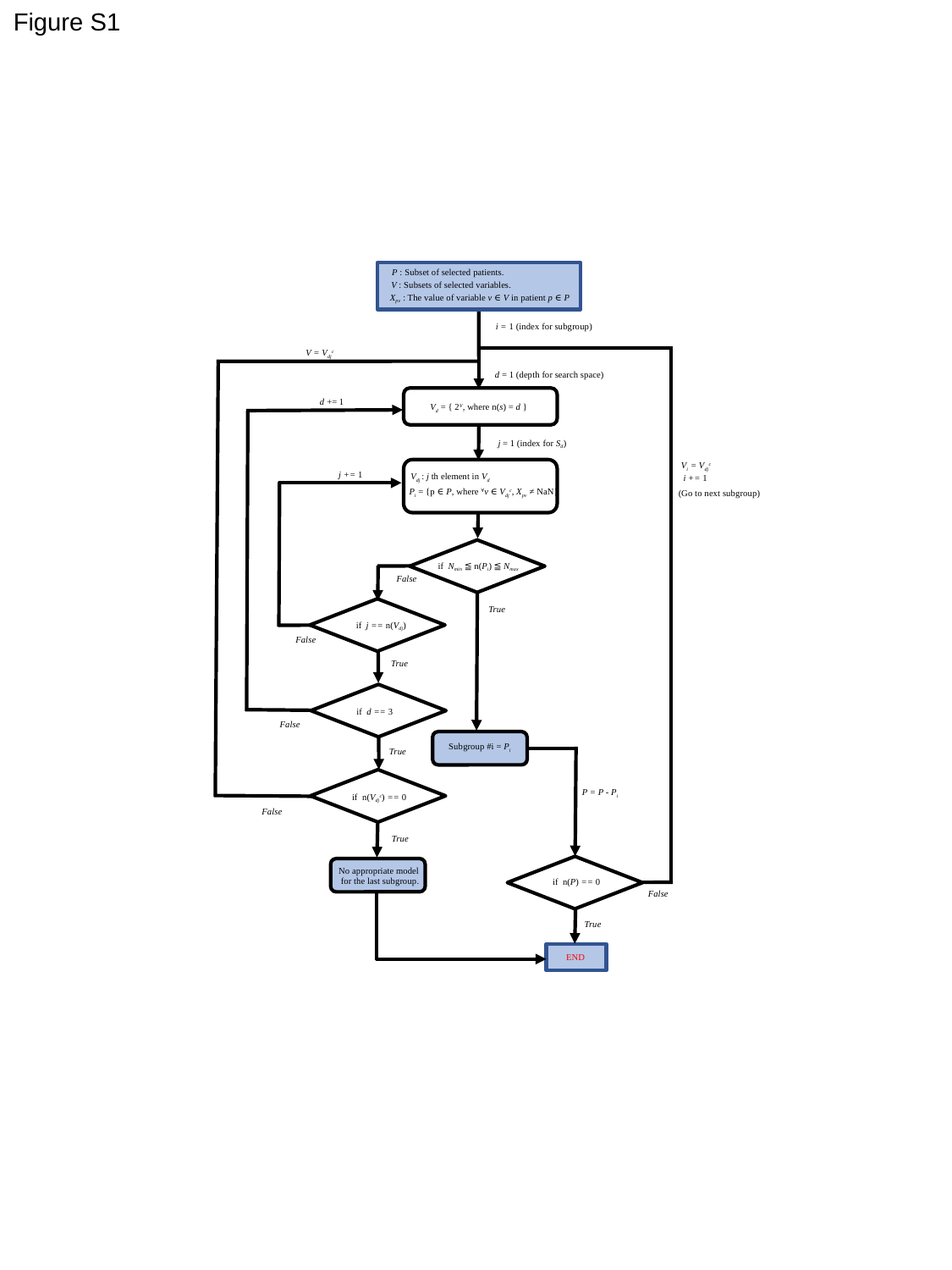

Figure S1
P : Subset of selected patients.
V : Subsets of selected variables.
X_pv_ : The value of variable v ∈ V in patient p ∈ P
i = 1 (index for subgroup)
V = V_dj_^c^
d = 1 (depth for search space)
V_d_ = { 2^V^, where n(s) = d }
d += 1
j = 1 (index for S_d_)
V_i_ = V_dj_^c^
V_dj_ : j th element in V_d_
P_i_ = {p ∈ P, where ^∀^v ∈ V_dj_^c^, X_pv_ ≠ NaN}
j += 1
i += 1
(Go to next subgroup)
if N_min_ ≦ n(P_i_) ≦ N_max_
False
True
if j == n(V_dj_)
False
True
if d == 3
False
Subgroup #i = P_i_
True
P = P - P_i_
if n(V_dj_^c^) == 0
False
True
No appropriate model
for the last subgroup.
if n(P) == 0
False
True
END

### Slide 2
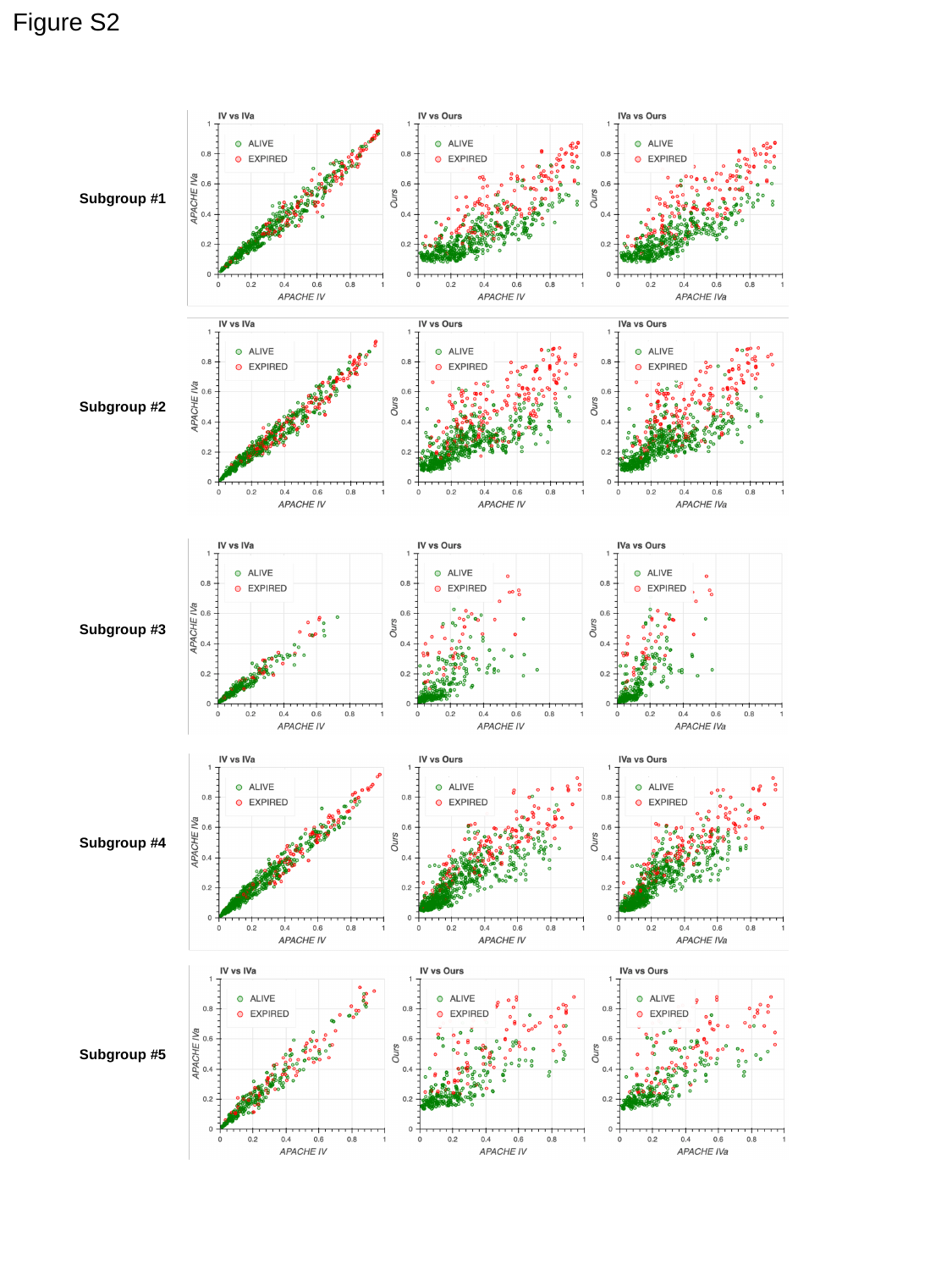

Figure S2
Subgroup #1
Subgroup #2
Subgroup #3
Subgroup #4
Subgroup #5

### Slide 3
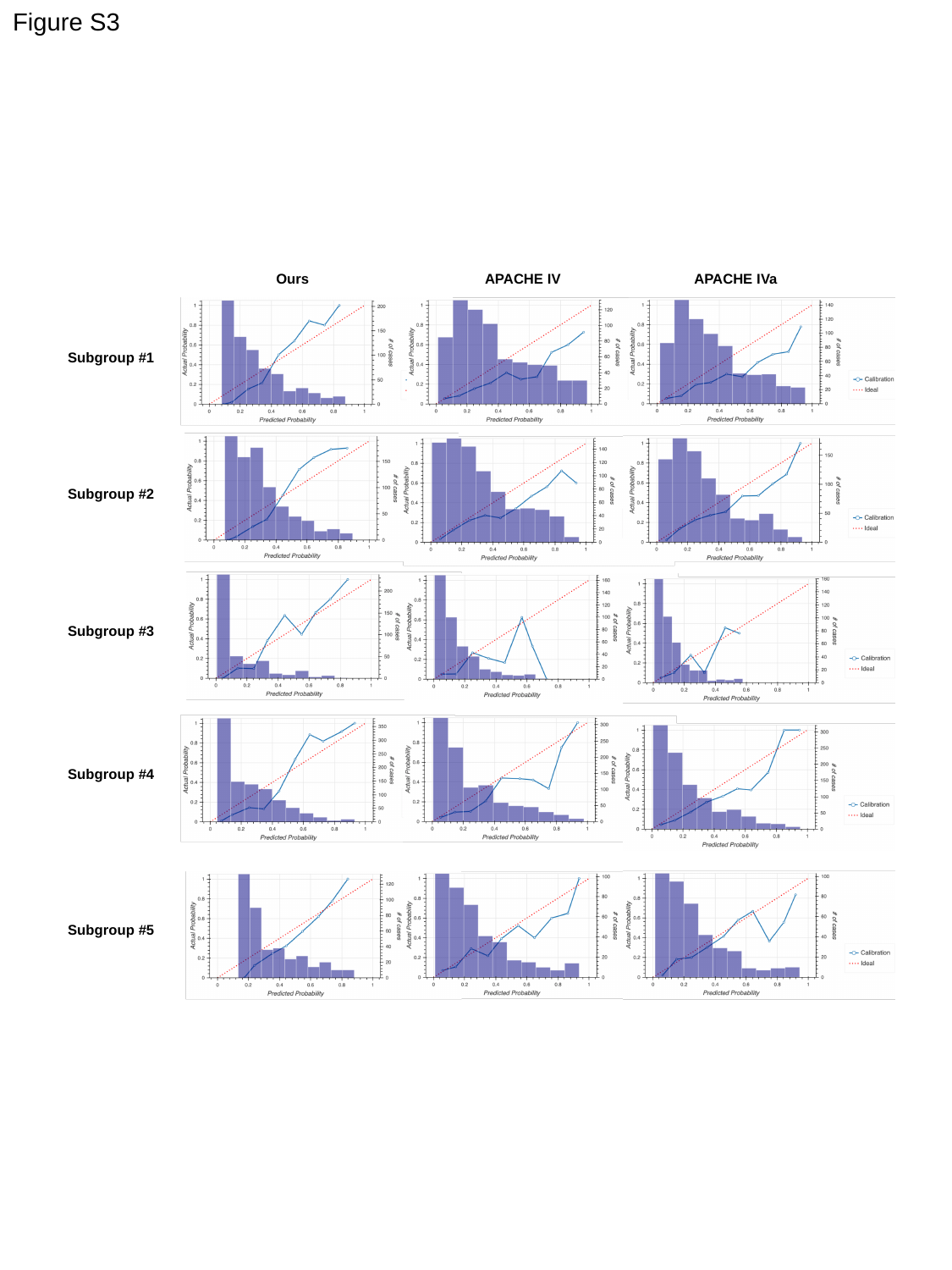

Figure S3
Ours
APACHE IV
APACHE IVa
Subgroup #1
Subgroup #2
Subgroup #3
Subgroup #4
Subgroup #5

### Slide 4
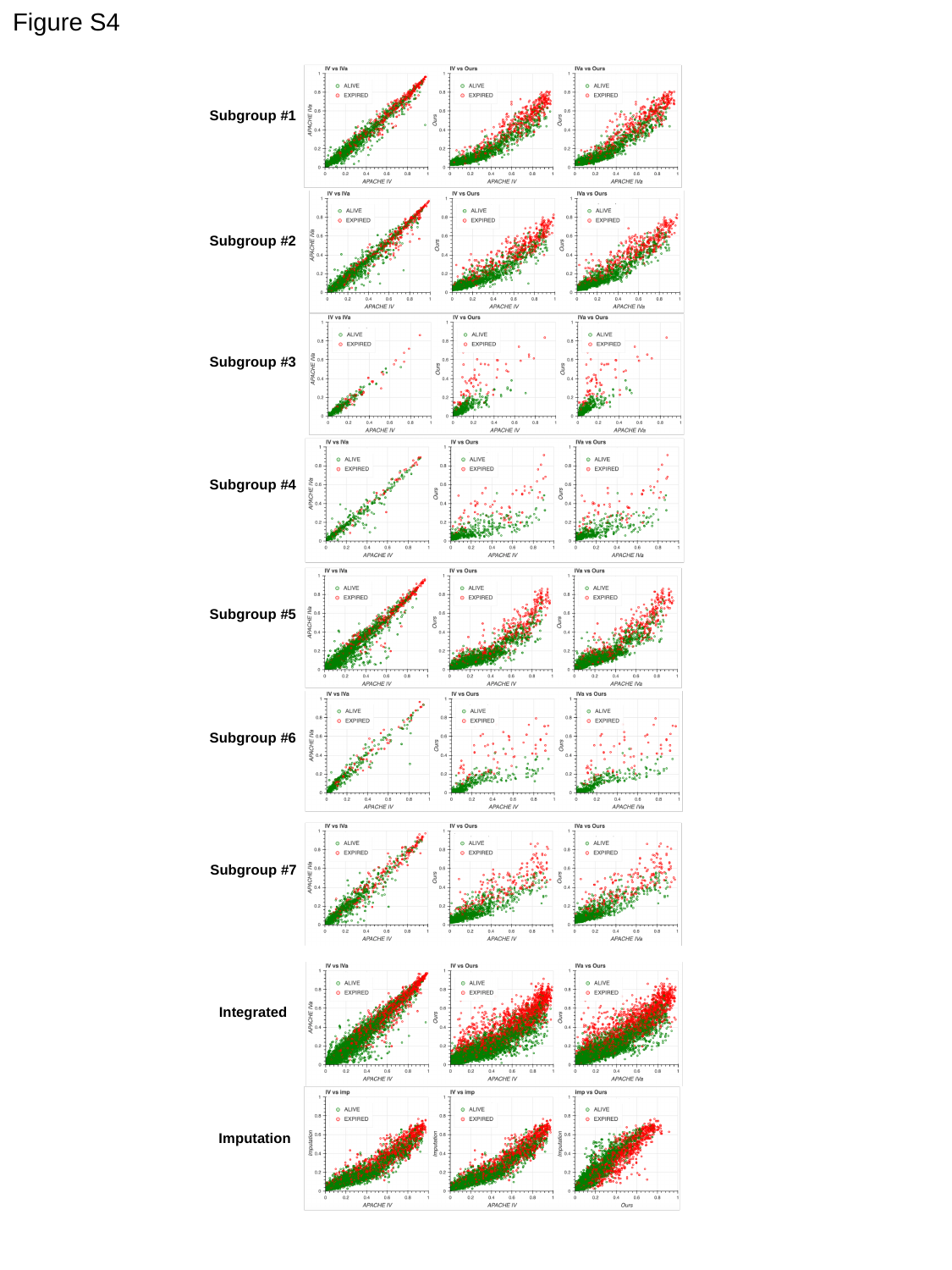

Figure S4
Subgroup #1
Subgroup #2
Subgroup #3
Subgroup #4
Subgroup #5
Subgroup #6
Subgroup #7
Integrated
Imputation

### Slide 5
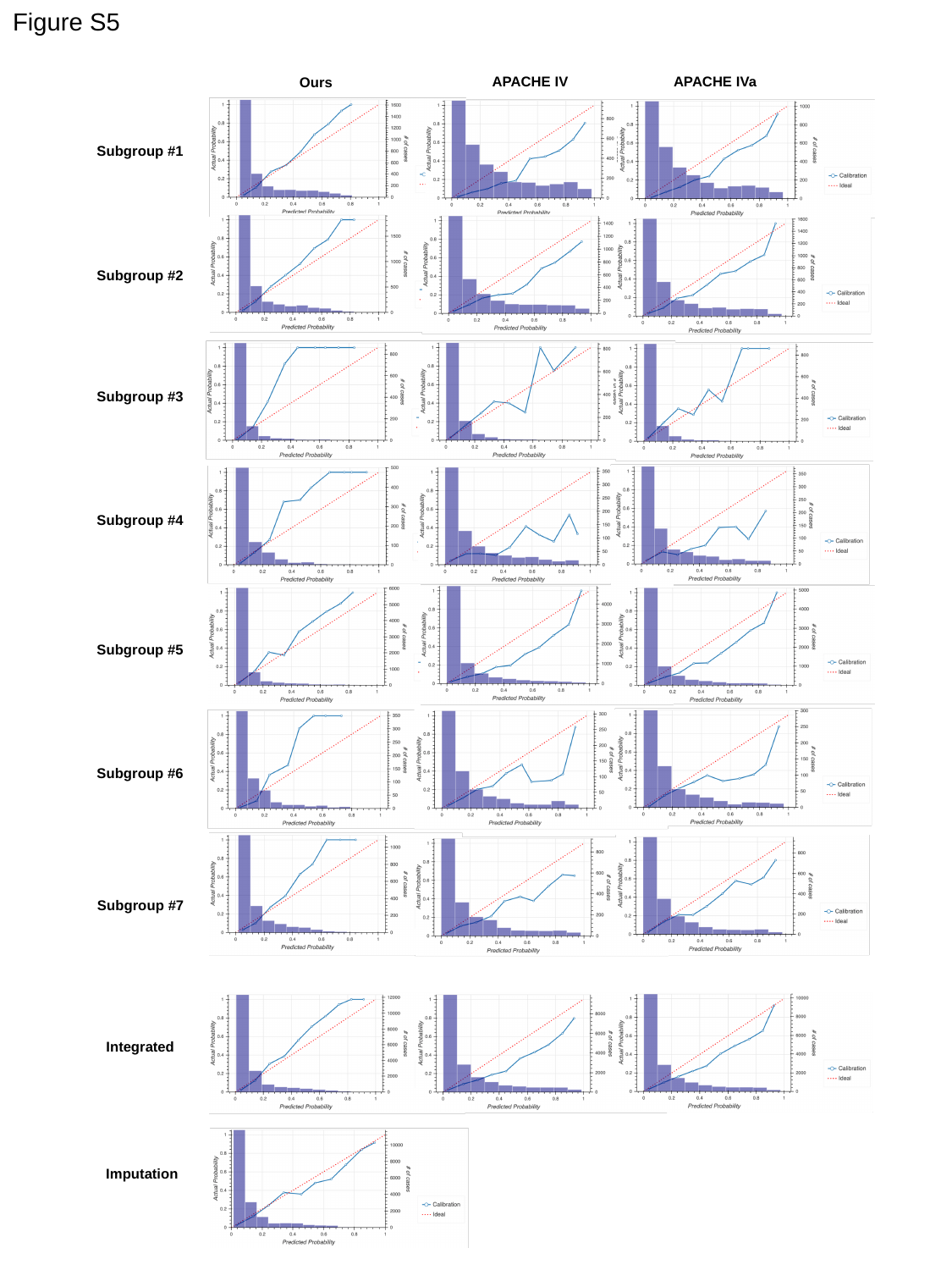

Figure S5
APACHE IV
APACHE IVa
Ours
Subgroup #1
Subgroup #2
Subgroup #3
Subgroup #4
Subgroup #5
Subgroup #6
Subgroup #7
Integrated
Imputation
